# Synoptic Reporting by Summarizing Cancer Pathology Reports using Large Language Models

**DOI:** 10.1101/2024.04.26.24306452

**Authors:** Sivaraman Rajaganapathy, Shaika Chowdhury, Vincent Buchner, Zhe He, Xiaoqian Jiang, Ping Yang, James R. Cerhan, Nansu Zong

## Abstract

**Background:** Synoptic reporting, the documenting of clinical information in a structured manner, is known to improve patient care by reducing errors, increasing readability, interoperability, and report completeness. Despite its advantages, manually synthesizing synoptic reports from narrative reports is expensive and error prone when the number of structured fields are many. While the recent revolutionary developments in Large Language Models (LLMs) have significantly advanced natural language processing, their potential for innovations in medicine is yet to be fully evaluated.

**Objectives:** In this study, we explore the strengths and challenges of utilizing the state-of-the-art language models in the automatic synthesis of synoptic reports.

**Materials and Methods:** We use a corpus of 7,774 cancer related, narrative pathology reports, which have annotated reference synoptic reports from Mayo Clinic EHR. Using these annotations as a reference, we reconfigure the state-of-the-art large language models, such as LLAMA-2, to generate the synoptic reports. Our annotated reference synoptic reports contain 22 unique data elements. To evaluate the accuracy of the reports generated by the LLMs, we use several metrics including the BERT F1 Score and verify our results by manual validation.

**Results:** We show that using fine-tuned LLAMA-2 models, we can obtain BERT Score F1 of 0.86 or higher across all data elements and BERT F1 scores of 0.94 or higher on over 50% (11 of 22) of the questions. The BERT F1 scores translate to average accuracies of 76% and as high as 81% for short clinical reports.

**Conclusions:** We demonstrate successful automatic synoptic report generation by fine-tuning large language models.

## 1. Introduction

Synoptic reporting is the process of formatting narrative pathology reports into a specific structured format [1]. The College of American Pathologists (CAP) has promoted the use of synoptic reporting since 1998, due to a number of advantages for various stakeholders in cancer diagnosis and treatment [1,2]. The standardized presentation, improved completeness, and conciseness enables clinicians to access pertinent information with greater ease and speed [1,3–5]. Through improve interoperability, synoptic reports also enable data registrars to collect and build large scale databases that are crucial for quality control, public health reporting, and research [2,6]. Most importantly, they encourage compliance to standards of care and accurate exchange of vital information, improving patient care and potentially patient outcomes [7,8].

The College of American Pathologists defines the synoptic reports as consisting of *data elements* and corresponding *element responses* [9] that capture the information present in an unorganized narrative text [6]. Documentation and administrative work has been shown to take 22% of a clinician’s work day on average [10]. The writing of reports in a highly engineered synoptic fashion, is known to further add to this high administrative demand on clinicians [6,11,12]. In addition, the transcription error rates have been positively correlated with the number of unique data elements required, implying a trade-off between detail and accuracy [1,3]. The standards for the minimum number and the type of data elements needed in a particular case is still evolving [4,6,8,9]. The conversion of existing narrative reports, while important for research, is yet another source of burden on clinicians and allied staff. To leverage the advantages while navigating these challenges necessitates automated synoptic reporting.

Existing work on automating synoptic reporting are limited in number and scope. Savova et al. [13] developed an open-source tool DeepPhe to extract cancer phenotype data from electronic health records (EHR). The techniques from open information extraction and artificial intelligence (AI) tools are combined in a heuristic manner for structurizing pathology reports in [14]. This proposed heuristic method has a primary disadvantage in that it is *ad-hoc* and requires the extensive redevelopment of the heuristic for pathology reports from a new source [14]. Lam et al. [15] describe a rule-based natural language processing (NLP) system to convert a semi-structured extensible markup language (XML) document into more structured tabular data. How such system can be adapted to generate tabular data from unstructured natural language reports instead of the semi-structured XML documents was not made clear. Mu and colleagues [16] demonstrated that a bidirectional encoder representations from transformers (BERT) trained using a binary relevance (BR) method can analyze textual pathology synopses and map patients to diagnostic keywords. The use of binary relevance, where semantic labels are transformed to multiple binary predictions is reported to have the disadvantage of ignoring label correlations [16]. Rule-based NLP was used by Tan et al. [17] to generate synoptic reports. While a high performance was reported, the authors report this was limited to data elements specific to breast cancer pathology reports that have been standardized at their institute [17]. It can be seen that the existing state-of-the-art NLP based methods for extracting structured data from unstructured data suffer from being limited in the types of reports they can analyse [17,18], have the tendency to lose important semantic information [14], often require semi-structured data [15], or require large amounts of expensive expert labeled data [18].

Large Language Models (LLMs), a recent advancement in artificial intelligence, offer a potential method to overcome the limitations of NLP based approach to synoptic reporting. LLMs large arrays of deep neural networks that are trained on massive corpora of text data [19]. The training of the models is typically done in a semi-supervised fashion, wherein, only a small amount of the data need to be adjudicated and annotated by human experts [20]. This enables LLMs to leverage the knowledge contained in large databases in a more cost-effective manner, compared to traditional NLP methods [21]. The training of large models with large corpora has been shown to result in emergent abilities – that is, an ability that is present in a larger model but is not present or could not be extrapolated from the abilities of smaller models [22]. These key advantages have triggered studies on evaluating the effectiveness of LLMs in the clinical setting, such as clinical text annotation [21], medical question and answering [23], and reasoning in the medical domain [24] – which form essential components of synoptic reporting. While large language models offer a promising path for automatic synoptic reporting, inaccuracies due to fabricated facts and unverified training data, lack of interpretability of the results, and potential risk of security breaches are some major challenges [20] that require careful consideration before their implementation.

In this work, we test the suitability of different pre-trained LLMs for the task of synoptic report synthesis. Further, we develop a training strategy to fine-tune and improve model performance for the task of automatic synoptic reporting. To measure the performance of the models, we use the state-of-the-art BERT F1 score [25], and device a method to translate the comparison metric to a more meaningful accuracy on the synoptic reporting task. To ensure that our results are robust to changes in the source and formatting of the reports, we validate our models on a sample of external data. We demonstrate a pathway for automating synoptic reporting from narrative cancer pathology reports using large language models while providing comprehensive insights into their strengths and weaknesses for this task.

## 2 Methods

### 2.1 Data

We use the Mayo Clinic’s enterprise clinical data warehouse, the Unified Data Platform (UDP) to access information on patients with cancer [26]. For this study, we digitally extracted 7,774 cancer pathology reports of 7,228 unique patients. Each report consists of four main components, which are:

1. <u>Gross description</u>: A *free text* record of a pathologist’s observations of the tissue sample at the macroscopic level [27].
2. <u>Preliminary frozen section consultation</u>: A *free text* record of a pathologist’s observations obtained through a special rapid analysis of a tissue sample that is frozen and reviewed during an ongoing surgery [27, 28].
3. <u>Diagnosis</u>: A *free text* record of the pathologist’s definitive diagnosis that integrates knowledge from the gross description, the optional frozen section consultation, and microscopic analysis.
4. <u>Synoptic report</u>: A *structured* summary created from the gross description, the preliminary frozen section consultation, and the final diagnosis [9]. These synoptic reports are in use for patient care at the Mayo Clinic, hence we treat these reports as the gold standard reference for model development and evaluation.

By *free text*, we refer to reports that are written in a natural unstructured or partially structured manner by the pathologist. From here on, we refer to the combination of the free text elements, the gross description, the preliminary frozen section consultation, and the diagnosis as the ‘*unstructured report*’. The structured synoptic report follows the format prescribed by the College of American Pathologists [9]. The synoptic report consists of several *data elements* and corresponding *element responses* (e.g., *Specimen Laterality: Right*). The exact data elements included in each report varies. For training, validation, and testing of our models, we have utilized the top 22 most frequently occurring data elements as show in **Table 1**. We use 80% of the data for training and 20% for testing, ensuring that no patient is common to both the training and the test sets.

**Table 1:**
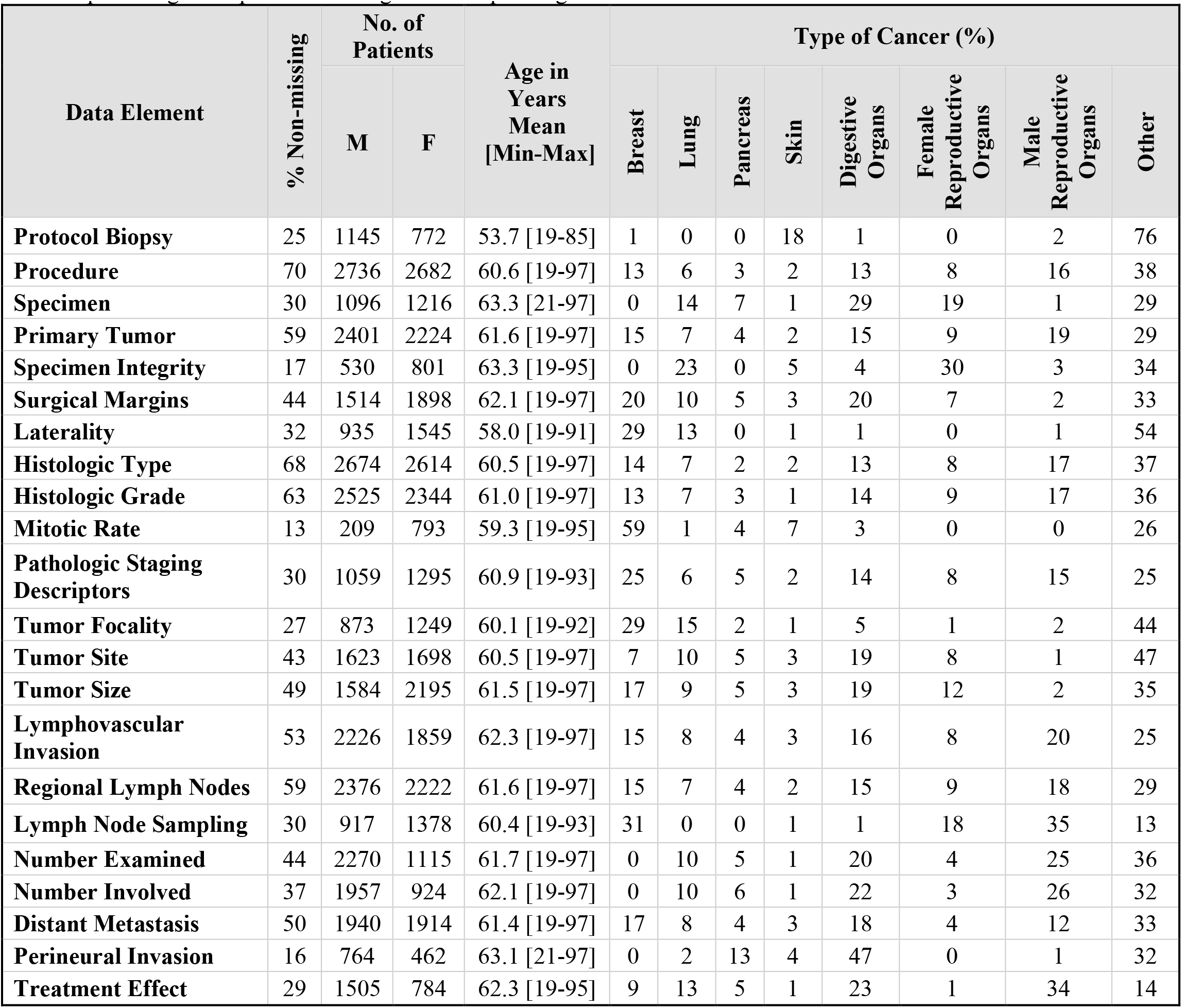
Summary of the 7,774 Mayo Clinic cancer pathology reports dataset. We show the top 22 most frequently reported data elements. The data elements listed do not occur in every report – the percent non-missing indicates the percentage of reports containing the corresponding data element.

### 2.2 Problem Definition

The synoptic reporting task we endeavor to automate is described as follows. Given an *unstructured report*, we utilize a large language model to generate a corresponding synoptic report with pre-specified data elements. We refer to the data element responses generated by a large language model as *‘estimated element responses*.*’* We refer to the data element responses filled in by the pathologist in the original data’s synoptic reports as *‘reference element responses*.*’* We evaluate the correctness of the estimated element responses by comparing with the reference element responses.

To automate the synoptic report synthesis using LLMs, we employ a prompt-based [30], element-by-element approach. An overview of this strategy is shown in **Figure 1**. We create an inference prompt that requests the model to find the response to a data element of interest. Such an inference prompt consists of an instruction segment, which indicates the data element of interest, along with a context segment, which is composed of the unstructured report. For each data element of the synoptic report, we create dedicated prompts by replacing the data element with the one of interest in the instruction segment. To generate an estimated element response using a large language model, we query it using the inference prompt. By sequentially querying a model for every data element using the inference prompts, we find the estimated element responses to each data element of the synoptic report. The same prompts are used for both pre-trained (zero-shot) and fine-tuned models. For fine-tuning a pre-trained model, we use training prompts. Training prompts are identical to inference prompts, except for one addition – the reference element response is added in the prompt. For fine-tuning, the model’s weights are updated such that its response matches the reference response.

**Figure 1:**
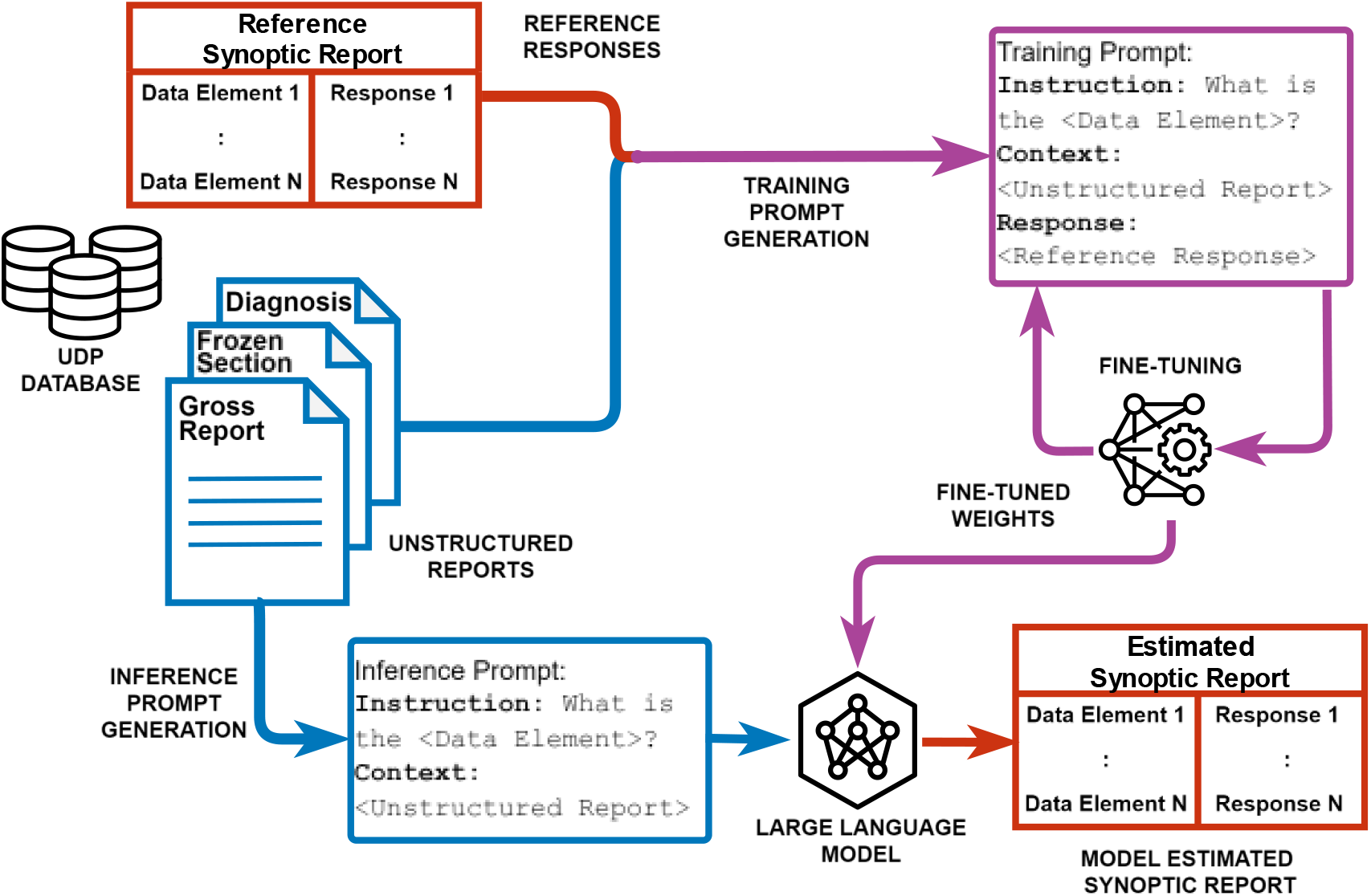
An overview of the large language model based automatic synoptic reporting. To generate a synoptic report from an unstructured pathology report, we take an element by element, prompt-based approach. We start by combining the gross report, frozen section report, and the diagnosis into a single unstructured report. We then generate an inference prompt, that includes an ‘instruction’ segment to query for a data element of interest and a ‘context’ segment that includes the unstructured report. The prompt is sent to the large language model whose response to the prompt is used to fill out the estimated element response of a synoptic report. The process is then repeated for other data elements. The same prompting strategy for inference is used for both pre-trained (zero-shot) and fine-tuned models. For fine-tuning a model, we use a training prompt, which is similar to an inference prompt, with the addition of a reference response. The model’s weights are updated to bring the model’s estimated response closer to the reference.

### 2.3 Models Used

We selected three classes of models for our study, the Bidirectional Encoder Representations from Transformers (BERT) [31], the Generative Pre-trained Transformer 2 (GPT-2) [32], and the Large Language Model Meta AI-2 (LLAMA-2) [33]. The exact variants of the model class used, and their relevant features are summarized in **Table 2**. The number of parameters roughly correspond to the size and complexity of the model. The maximum input length is the limit on the size of a block of text a model can process. This input length is measured in tokens, where 1 token is approximately 4 characters in length. All the models we use are *pre-trained*, wherein the models have been previously trained on a large corpus of publicly available, non-specialized textual data [31–33]. We use the offline, fully accessible versions of these models offered through the Hugging Face Transformers library in Python [34].

**Table 2:** A summary of the model classes used in this study and their relevant features. Model variant refers to the exact variant of the model used from a selection available. The number of parameters represents the complexity and size of the model. The maximum input length is the maximum number of tokens (1 token is approximately 4 characters) in an input text the model can process at a time. The training data is the dataset that was used to pre-train the model.

| Model Class | Model Variant Used | Number of Parameters | Maximum Input Length (tokens) | Data Used for Pre-Training |
| --- | --- | --- | --- | --- |
| BERT | BERT large model (uncased) | 334M | 512 | Dataset of 11,038 unpublished books and English Wikipedia [31]. |
| GPT-2 | GPT-2 | 124M | 1024 | A proprietary dataset created by mining the web for heuristically determined high quality text [32]. |
| LLAMA-2 | LLAMA-2-7b | 7000M | 4096 | A proprietary dataset of 2 trillion tokens created from publicly available sources without sources rich in personal and private information [33]. |

**Table 3:** The grouping of the data elements based on the type of reasoning needed by a large language model to produce the response. Here we report the total fraction of reports in which the reference element response (or part of the reference) is present. Furthermore, we also show how many times the reference element response (or part of the reference) is present in the unstructured report.

| Data Element | Reference in Unstructured Report (%) | Number of Occurrences (Median) | Reasoning Type |
| --- | --- | --- | --- |
| SURGICAL MARGINS | 100.0 | 19 | Inductive |
| SPECIMEN | 96.3 | 18 | Inductive |
| LATERALITY | 96.1 | 12 | Inductive |
| HISTOLOGIC TYPE | 91.4 | 4 | Inductive |
| LYMPH NODE SAMPLING | 91.2 | 19 | Inductive |
| TUMOR SIZE | 87.2 | 14 | Inductive |
| TUMOR SITE | 83.9 | 7 | Inductive |
| PROCEDURE | 79.7 | 3 | Inductive |
| TREATMENT EFFECT | 78.6 | 2 | Inductive |
| TUMOR FOCALITY | 77.2 | 14 | Abductive |
| HISTOLOGIC GRADE | 73.0 | 2 | Abductive |
| PERINEURAL INVASION | 66.9 | 1 | Deductive |
| DISTANT METASTASIS | 53.6 | 1 | Deductive |
| LYMPHOVASCULAR INVASION | 50.8 | 1 | Deductive |
| MITOTIC RATE | 41.5 | 0 | Abductive |
| PATHOLOGIC STAGING DESCRIPTORS | 38.9 | 0 | Deductive |
| SPECIMEN INTEGRITY | 28.3 | 0 | Deductive |
| PRIMARY TUMOR | 25.0 | 0 | Deductive |
| NUMBER INVOLVED | 11.5 | 0 | Abductive |
| NUMBER EXAMINED | 4.9 | 0 | Abductive |
| REGIONAL LYMPH NODES | 3.9 | 0 | Deductive |
| PROTOCOL BIOPSY | 2.2 | 0 | Deductive |

### 2.4 Evaluation Strategy

To evaluate the performance of the models, we compare the estimated element responses against the reference data element responses in the synoptic reports written by the pathologists. While one strategy for this comparison could be simple string matching, this leads to incorrect assessment. For example, the estimated response ‘*Invasive adenocarcinoma*’ compared against a reference response of *‘Adenocarcinoma is Invasive’* will lead to a poor score if done by exact string matching. This necessitates a more robust strategy to evaluate the model’s estimated responses. We utilize the BERT F1 score [25] to measure model performances. To obtain an accuracy that corresponds to human evaluation, we determine a threshold by comparing the BERT F1 scores against manual evaluation on a test set composed of 100 unstructured reports for each of the 22 data elements (see Appendix A **Figure 7**). We use a threshold of 0.85, i.e. a BERT F1 > 0.85 is needed for estimated responses to be considered correct.

### 2.5 Description of Experiments

Here we describe our experiments to determine efficacy of LLMs in automating the synoptic reporting task.

#### 2.5.1 Pre-trained vs Fine-tuned models

The models we selected (BERT, GPT-2, and LLAMA-2) to study have been pre-trained on large corpora of unstructured text data. The dataset used for the pre-training is unspecialized and is likely not suited for the highly specialized task of synoptic reporting. While prompting [30] has been shown to enable LLMs to perform specialized downstream tasks, only moderate performance has been reported for annotating medical texts [35]. Therefore, we *fine-tune* [36] the models, wherein the model parameters (or a fraction of the model parameters) are updated through training on relevant medical text, to improve performance on the downstream task [37]. We use the Transformers library [34] to perform the model fine-tuning for BERT and GPT-2. For fine-tuning the much larger LLAMA-2 models, we use the quantized low-rank adaptation (QLoRA), a parameter efficient fine-tuning strategy [38]. We use zero-shot prompting, where only the query and context are sent to the model without any prior examples [30]. We use the zero-shot method to efficiently utilize the limited input token sizes. We use identical prompts for both pre-trained and fine-tuned models and compare them on the same test dataset.

#### 2.5.2 Handling Large Reports

It can be seen from **Table 2** that the large language models have strict limits on the number of words they can process in an input. A significant number (∼56% for LLAMA-2) of our unstructured reports exceed the input limits of even the LLAMA-2 model with a capacity of 4096 tokens. We handle such large reports by chunking them into smaller contiguous pieces so that each model has an opportunity to scan the whole unstructured report. The best estimated data response amongst the estimates obtained from each piece is selected for evaluation. We focus the analysis on the best performing model (LLAMA-2) from this experiment onwards.

#### 2.5.3 Effect of the Size of Fine-Tuning Data and Model Size

Here we aim to quantify the amount improvement capacity left in the fine-tuned models. We do this by varying the dataset size used for fine-tuning and reporting the test performances of the partially fine-tuned models. We also test two variants of the LLAMA-2 model – the 7 billion and 13 billion parameter variants.

#### 2.5.4 Inductive, Abductive, and Deductive Reasoning

The reasoning capabilities of large language models are still under investigation [39,40]. We stratify the performance of our best performing fine-tuned model based on *inductive, abductive, and deductive* reasoning [40]. We accomplish this by measuring model performances on the data elements grouped into the following three categories:

a. Inductive elements: are element responses that involve generalizing based on specific observations.
b. Abductive elements: are element responses that involve selecting one evidence from multiple that best fits a set of observations.
c. Deductive elements: are element responses that involve deriving a conclusion from given premises of principles.

#### 2.5.5 External Validation

Sushil et al. [35] provide a sampling of oncology reports that have been curated by experts, for the purpose of testing the performance of language models. We use this data for external validation. The curated dataset consists of 40 deidentified breast and pancreatic cancer progress notes. A team of medical experts have labeled the unstructured notes at a fine-grained level using BRAT, an open-source annotation software [35]. Three categories of labels have been given to the text, which include entities, attributes of the entities, and relationships between the entities. We reviewed these labels and determined that only 5 labels under the entities category correspond with 5 of 22 data elements we have considered in our analysis. These are Histologic Type, Laterality, Tumor Site, Primary Tumor, and Procedure. We use our models to extract the responses to these 5 data elements from their reports and compare the model responses to the expert-annotated labels.

## 3 Results

In this section, we present the results in detail.

### 3.1 Performance improvement with Fine-tuning

Figure 2. compares the performance of the pre-trained and fine-tuned models on estimating the data element responses. **Figure 2 (a)** shows a summary of the BERT F1 scores for the test set composed of 22 data elements. Fine-tuning improves median of the score, with LLAMA-2 having the largest increase (Median F1 score from 0.68 to 1). The inter quartile range (IQR, which is the difference between the 75th and 25th percentiles of the data) [41] of the fine-tuned LLAMA-2 model increases from 0.04 to 0.13, indicating that the model has a wide range of performances depending on the data element type. This is clarified in **Figure 2 (b)**, which shows the detailed stratified performance of the pre-trained and fine-tuned models on each data element tested. The numbers inset in the heat map show the mean of the F1 score. It can be seen that the range of scores for the pre-trained LLAMA-2 varies from 0.67 to 0.70, while the fine-tuned version ranges from 0.81 (for Pathologic Staging Descriptors) to 1 (for Protocol Biopsy).

### 3.2 Effect of Report Size

When encountering unstructured reports whose length exceeds the input limit of an LLM, we are forced to split the report into contiguous pieces. This reduces the field of view of the model a fraction of the unstructured report. In **Figure 3**, we show the performance degradation of fine-tuned LLAMA-2 due to the reduction in the field of view. When the complete unstructured report fits in the field of view of the model, its performance is improved: the median BERT F1 scores increase to 1 (IQR = 0.09) from 0.97 (IQR = 0.09). The projected accuracy scores show an improvement of 12 percentage points (81% for short reports vs 69% for long reports).

**Figure 2.**
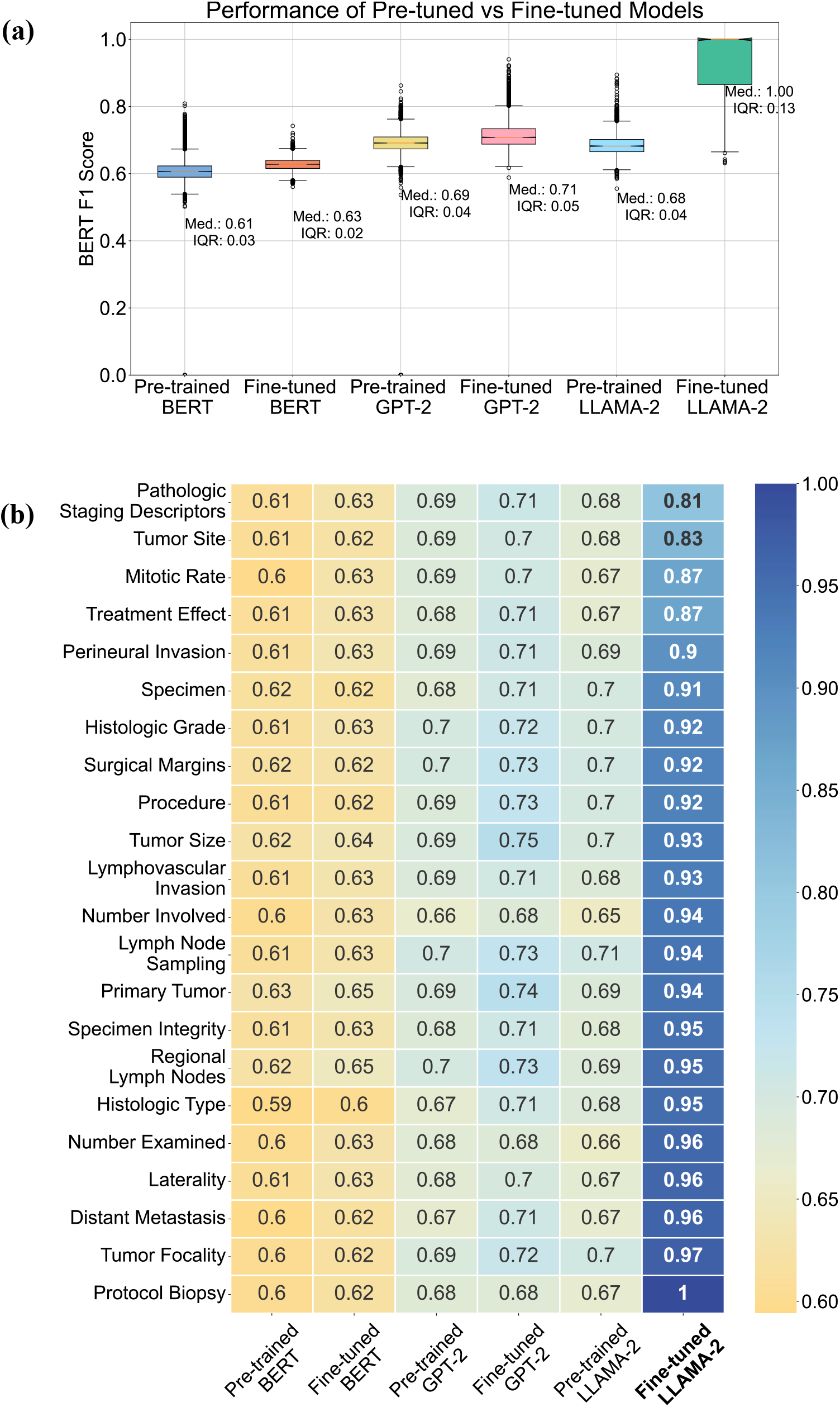
**(a)** Performance comparison of pre-trained and fine-tuned language models on the synoptic reporting task. A BERT F1 Score of 1 indicates a perfect match between the model’s output and the reference answer. The median and the Inter Quartile Range (IQR) [41]are reported in the plots. **(b)** Performance of the pre-trained and fine-tuned language models, segregated by the question. The numbers inset in the heat map show the mean BERT-F1 score.

**Figure 3.**
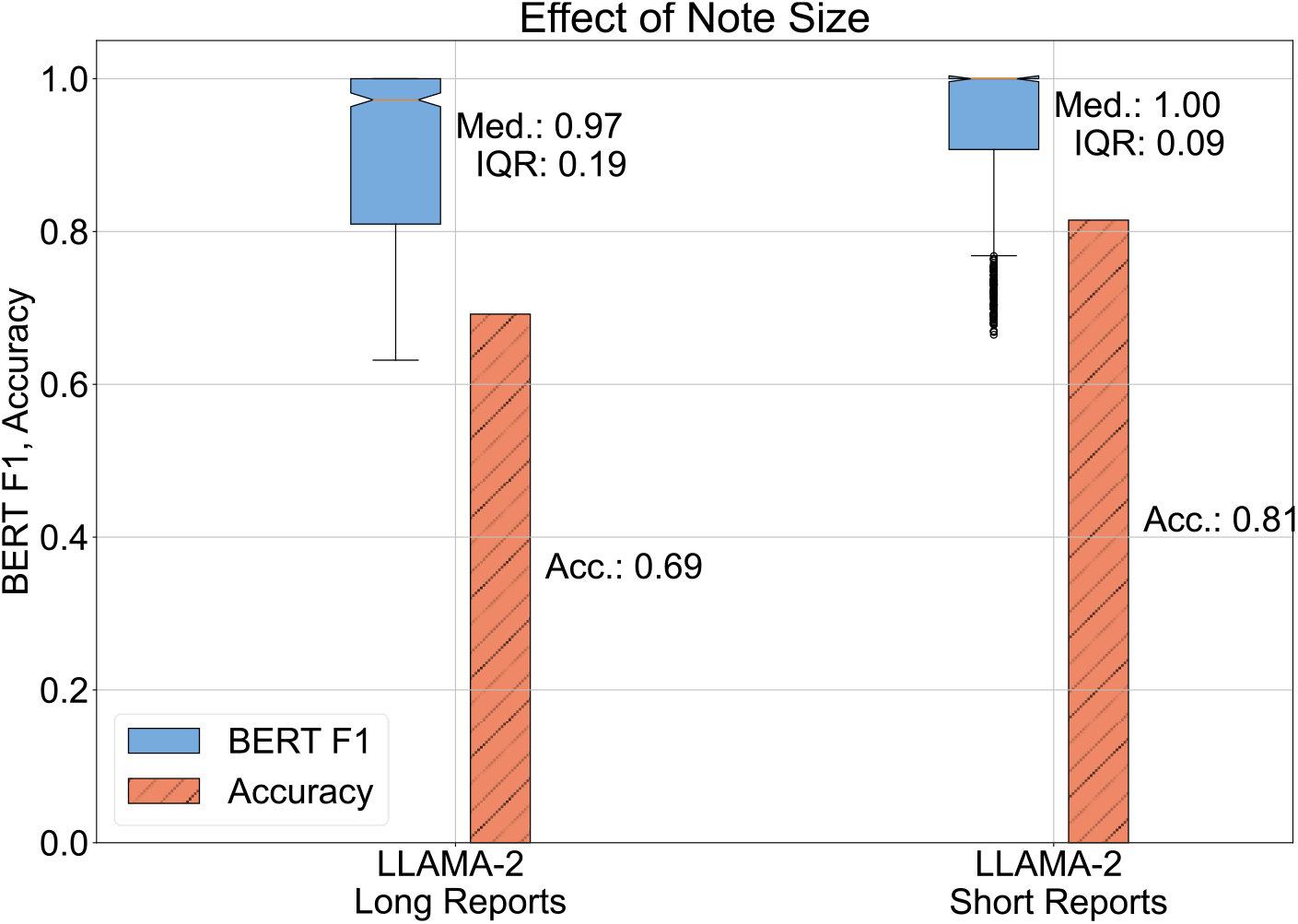
Performance differences in the fine-tuned LLAMA-2 model with variation in the size of the clinical notes. Long reports are clinical notes whose size (in no. of words) exceeds the token limit (4096), whereas short reports are clinical notes whose size (in no. of words) is less than the model’s token limit. Long reports are broken into smaller pieces to fit the model’s token limit, leading to its inability to answer questions accurately, leading to a reduction in performance. The projected accuracy is computed using a threshold of BERT F1 > 0.85 to closely align with manual analysis.

### 3.3 Effect of the Size of Training Data and Model Size

In **Figure 4**, we present the gain in performance we can achieve if the size of the training data is increased. From **Figure 4 (a)** we observe a small improvement in the performance, with the projected accuracy changing from 0.70 to 0.76 when the size of the training data is increased from 25% to 100%. An increase in the parameter size by roughly 50% from 7 billion to 13 billion does not produce a corresponding improvement in performance: the 13 billion parameter variant has accuracies that are 1 to 2 percent points higher than its 7 billion parameter variant.

**Figure 4.**
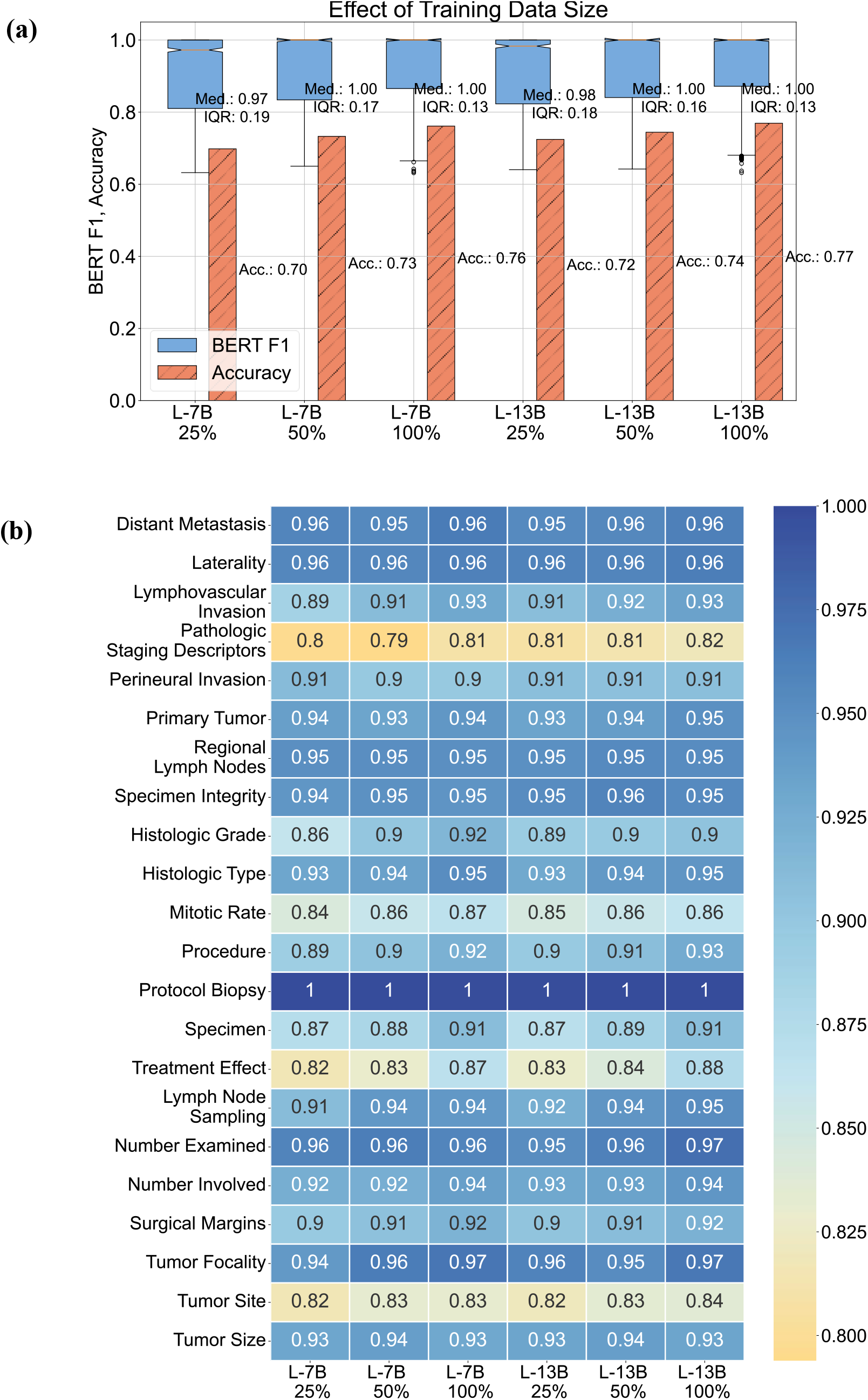
**(a)** Performance comparison of fine-tuned LLAMA-2 models on the synoptic reporting task as the size of the training data is increased. Two variants of the LLAMA-2 model are tested, the 7 billion parameter version (L-7B) and the 13 billion parameter version (L-13B). **(b)** Stratified performance scores for each data element. The numbers inset in the heat map show the BERT-F1 score.

### 3.4 Inductive, Abductive, and Deductive Reasoning

We compare the performances of the LLAMA-2 model on different reasoning tasks in **Figure 5**. We observe a reduction in accuracy for inductive reasoning tasks, which have a projected accuracy of 0.73 vs those for deductive reasoning tasks with an accuracy of 0.79.

**Figure 5.**
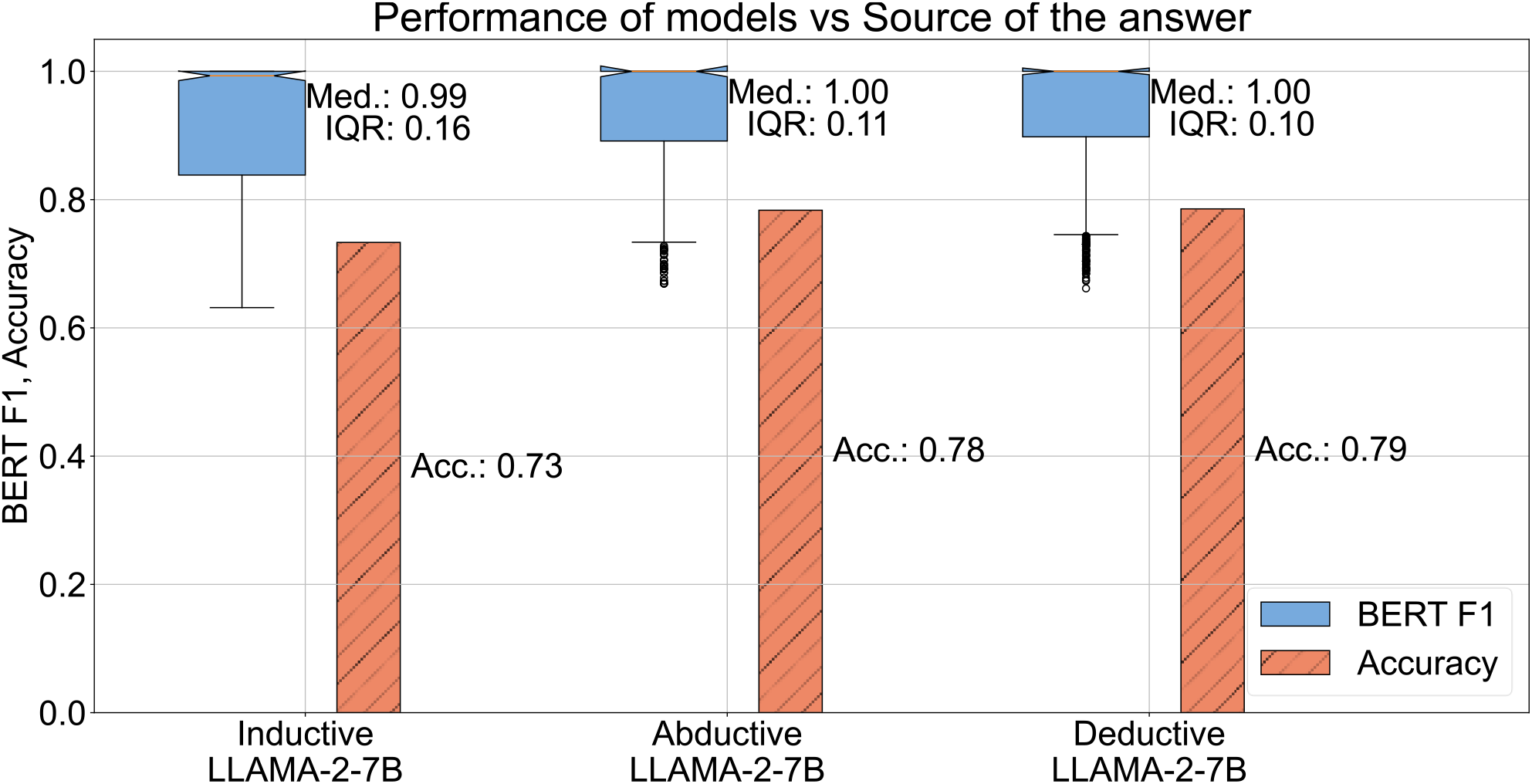
Performance differences of the fine-tuned LLAMA-2 model on questions whose answers are categorized into three different groups based on the type of reasoning needed for producing an element response.

### 3.5 External Validation

In **Figure 6**, we compare the performances of the pre-trained and fine-tuned LLAMA-2 7B models on the external, expert-annotated data from [35]. We compare the responses given by the models to identical prompts, for 5 data elements that correspond to the labels provided in the external dataset. We compare the responses against the labels manually and report the accuracies. The fine-tuned LLAMA-2 model described here is the same as model described in earlier parts of this study – it has only been fine-tuned on our data; it has never been shown any examples from the external data. We observe that the fine-tuned LLAMA 2 outperforms the pre-trained version. The fine-tuned model has good performance on determining the histology (97.2% accuracy) and tumor laterality (84.4% accuracy). The performance is reduced for tumor site, primary tumor, and procedure (67.5%, 46.2%, and 41.0% respectively). The most probable reasons for the reduced performance are likely due to differences in the nature of the reporting styles and the larger size of the external data’s unstructured reports (the external data’s unstructured reports are 5-10x larger than ours).

**Figure 6.**
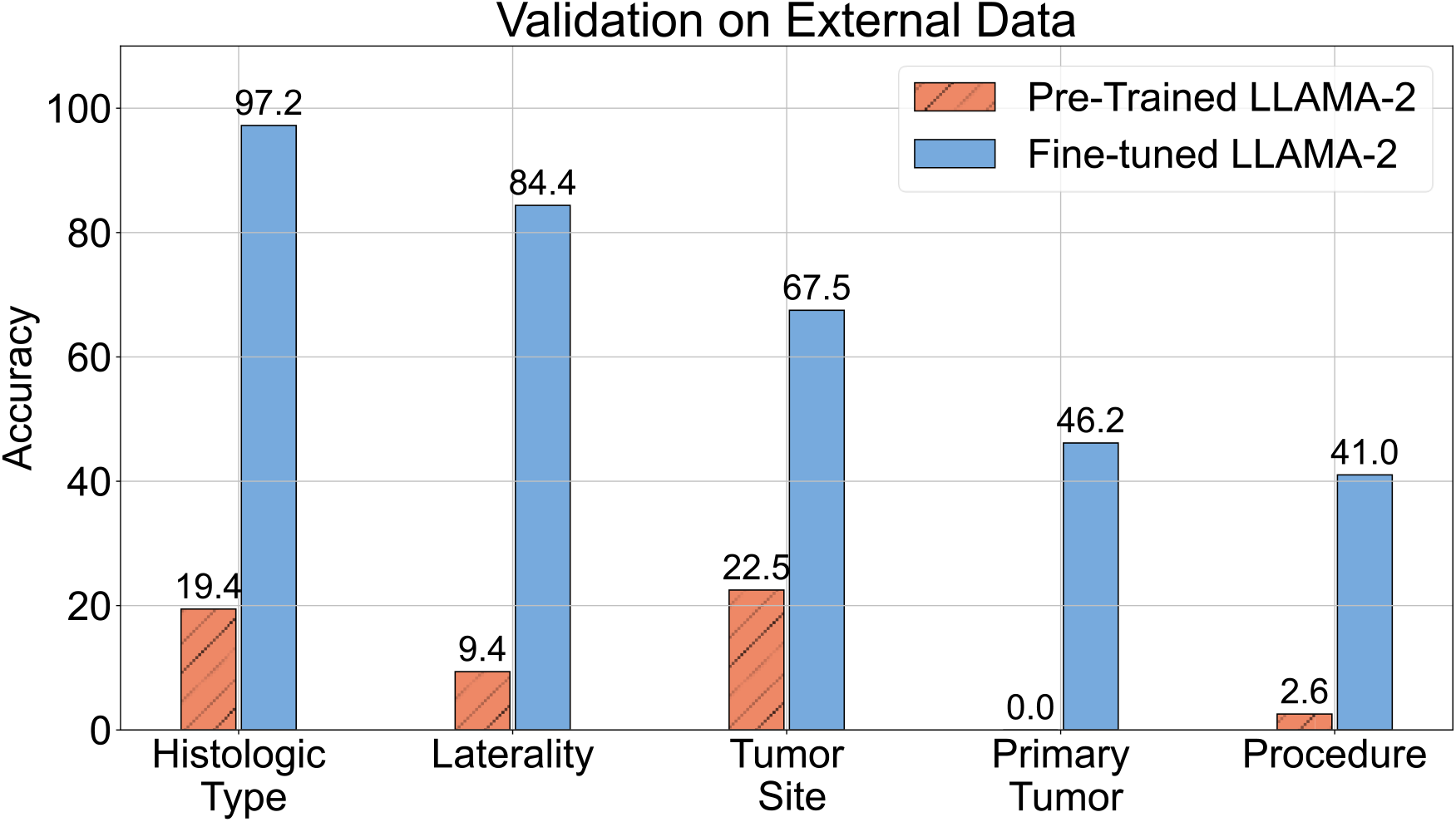
Performance of the pre-trained and fine-tuned LLAMA-2 7B model (best performing model) on an external dataset in Sushil et al. [35]. We test the ability of the two models to extract 5 data elements which correspond to the labels provided in the external dataset. The accuracy reported here is obtained by manual comparison of the labels provided in [35] against the model responses. Identical prompts were used for both the pre-trained and fine-tuned models. It can be seen that the fine-tuned model outperforms the pre-trained model by a large margin. *Note: Fine-tuned LLAMA-2 has only been fine-tuned on our data, it has not seen examples from the external data*.

**Figure 7:**
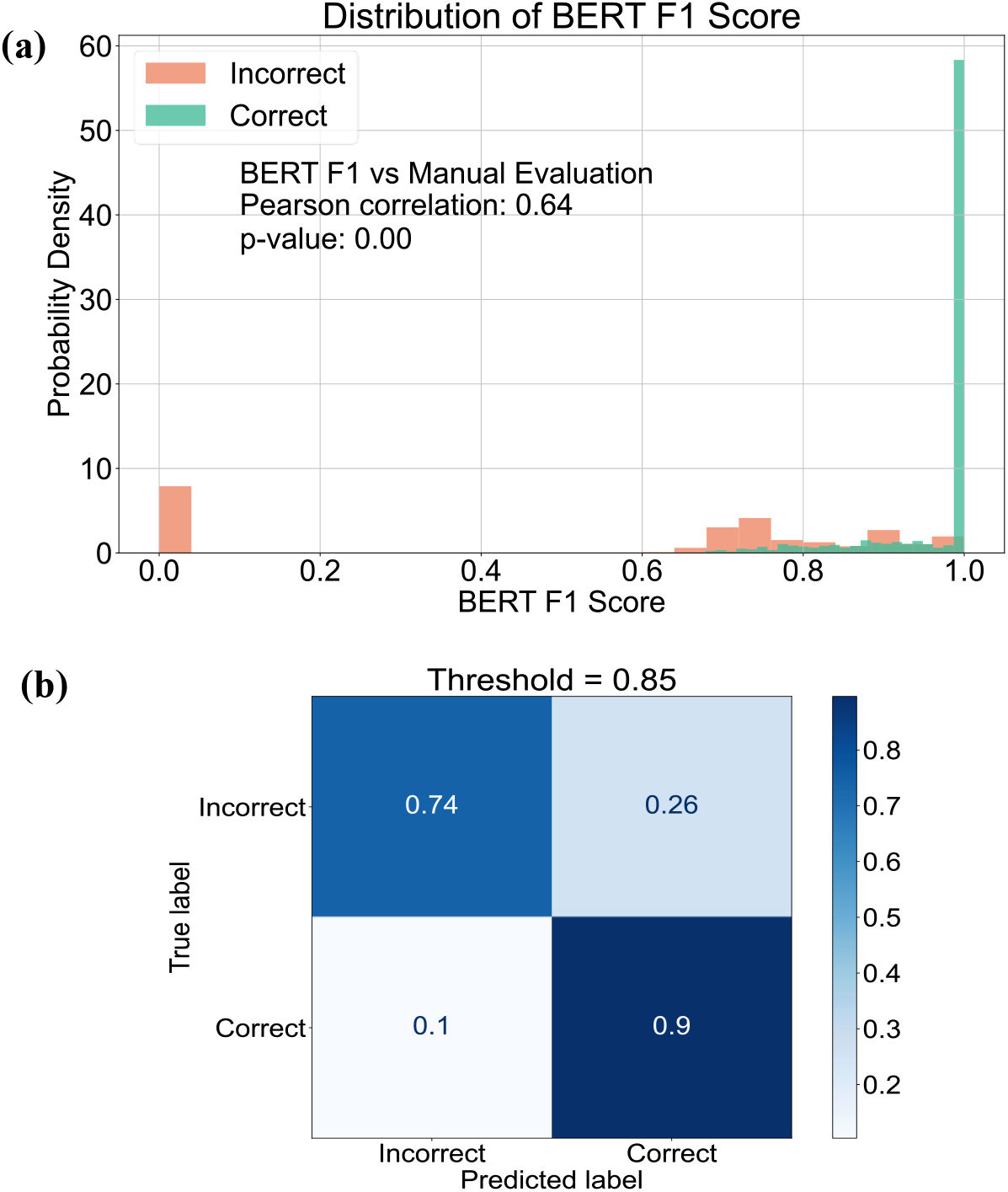
Evaluation strategy - projected accuracy from BERT F1 Score. In (a), we show the distribution of BERT F1 scores obtained by comparing the reference element responses and the estimated element responses for the LLAMA-2-7B model. A test set of 100 examples for 22 data elements were used. The correctness and incorrectness of a response was determined by manual comparison. The Pearson correlation between the manual score and BERT F1 is 0.64 and the p-value of 0 indicates the correlation is significant. (b) The confusion matrix for BERT F1 score based determination of the correctness of a response – a threshold of 0.85 is used in this study.

## 4. Discussion

In this study we showcase automatic synoptic reporting using large language models that have been pre-trained on large datasets of non-specialized unstructured data. We use an element-by-element prompting strategy to achieve this. Prompting the pre-trained models does not yield estimated element responses that are very accurate, yielding BERT F1 scores ranging from 0.61 to 0.68 (see **Figure 2 (a)**). Neither the size of the pre-trained model nor its complexity plays a sizable role in improving the performance with BERT, GPT-2, and LLAMA-2, yielding similar BERT F1 scores of 0.61, 0.69, and 0.68 respectively. Fine-tuning was observed to improve the performance, with greater improvement obtained in proportion to the size of the model. Indeed, the LLAMA-2 variant with 7 billion parameters after fine-tuning is able to achieve median BERT F1 scores of 1.00 (IQR = 0.13) from 0.68 (IQR = 0.04) for its pre-trained variant, as shown in **Figure 2 (a)**. This observation of a sudden significant improvement as the model size increases by 2 orders of magnitude corresponds well with the property of emergent behavior in LLMs [22].

What do the BERT F1 scores imply for the task of synoptic reporting? We show that the BERT F1 correlates well with accuracy (Pearson coefficient = 0.64) and project them to accuracies using a threshold of 0.85. This projected accuracy from BERT F1 aligns with manual evaluation (see Appendix A, **Figure 7**). Using this technique, we observe that with fine-tuning, we can achieve an average accuracy of 76% using the 7 billion parameter variant of LLAMA-2 (see **Figure 4 (a)**). In addition, we observe an improvement in performance with additional training data in **Figure 4 (a)**, indicating there is more room for improvement with increases in training data size. Our results indicate that when the size of the unstructured notes exceeds the input capacity of a model, the performance reduces. Thus, engineering the capability to handle reports that exceed the model input token limits has the potential to improve performances (81% from 69%, as seen in **Figure 3**). Through external validation on set of expert-labeled oncology reports as shown in **Figure 6**, we note that fine-tuning for the synoptic reporting task provides a significant advantage over using pre-trained models. We observe that even on unseen external data, with significantly larger reports (5-10x the internal training data), the performance of the fine-tuned model can be high (84.4 – 97.2% accuracy for Laterality and Histologic Type in **Figure 6**), showcasing the potential of fine-tuned LLMs. We acknowledge the following limitations to ensure the scope of our study is well defined. First, we did not engineer any strategy to handle reports whose sizes exceed the input token limit of an LLM so that the performance did not suffer. Second, we restricted our attention to generating element responses when a set of data elements have been prespecified. A more comprehensive automation strategy would be to generate a list of appropriate data elements when an unstructured note is presented. An alternative is to engineer a look-up table based list of data elements using the templates from the College of American Pathologists [9]. This however, needs an effective cancer type detection system from the unstructured reports. Third, due to the source of the data used, our study and observations are restricted to the writing styles utilized by clinicians at the Mayo Clinic. Expanding the dataset to include synoptic reports from other hospital systems and from clinicians whose first language is not English should improve robustness to writing style differences. Fourth, we used data that is used in practice as-is with no additional expert annotation as has been done by Sushil et al. [35] for creating their testing benchmark dataset. The lack of such expensive expert manual annotation restricts the resolution with which we can analyse the results. Fifth, our study uses EHRs linked to cancer. Automated generation of synoptic reports for diseases other than cancers is beyond the scope of this study.

## 5 Conclusions & Future work

We have demonstrated that using a sufficiently large language model, when fine-tuned appropriately, is able to estimate the element responses of a synoptic report with high average accuracy of up to 77%. We further validated our models on external expert-labeled data, where we showcase similar performance on select cases, with graceful reduction in performances in relation to pre-trained models. We believe that this capability effectively demonstrates that the use of fine-tuned LLMs is an effective strategy to automate synoptic reporting. We utilize offline, fully accessible large language models to complete our tasks. This enables us to modify the model architecture and potentially share the model weights for other lateral tasks. The offline models can also be kept secure within an organization to ensure patient privacy and improved security in a cost-effective manner. Handling large reports, either via summarization or via a vote-based estimate, needs to be investigated in the future. We have shown that this investigation has the potential to improve average accuracies up to 12 percentage points. Our work in this study assumes that the user queries the models with a set of desired data elements. Given an unstructured report, having an LLM determine the essential data elements (same as would be recommended by a pathologist), provides a more comprehensive and automated solution.

## Data Availability

Due to Mayo Clinic policy and to protect privacy, the raw electronics health records data cannot be shared. The data presented in the figures and the code used to train the models is available here: https://github.com/bioIKEA/SynopsisGPT

## Contributions

The study was conceptualized by NZ. NZ was responsible for extracting the raw data. SR was responsible for data post processing, developing the models, conducting the experiments, and preparing the data visualization. NZ and SR worked on the draft with other authors providing evaluation and feedback. VB contributed to integrated system development. All authors collectively took ultimate responsibility for deciding to submit the report for publication and committed to being accountable for all aspects of the work, ensuring that any concerns about accuracy or integrity were adequately addressed and resolved. NZ supervised and administered the study.

## Acknowledgments

The authors thank Andrew Wen, M.S., for assisting with the usage of the Mayo Clinic Unified Data Platform.

## Data Permissions

The data usage was approved by a Mayo Clinic Institute Review Board (IRB).

## Appendix A Evaluation Strategy

## References

1 Renshaw AA, Mena-Allauca M, Gould EW, et al. Synoptic Reporting: Evidence-Based Review and Future Directions. JCO Clinical Cancer Informatics. 2018;1–9.

2 Valverde L D, Reznichek RC, Torres SM. Establishing Synoptic Cancer Pathology Reporting in Low- and Middle-Income Countries: A Nicaraguan Experience. JCO Glob Oncol. 2022;8:e1900343.

3 Renshaw AA, Gould EW. Comparison of Accuracy and Speed of Information Identification by Nonpathologists in Synoptic Reports With Different Formats. Archives of Pathology & Laboratory Medicine. 2016;141:418–22.

4 Strickland-Marmol LB, Muro-Cacho CA, Barnett SD, et al. College of American Pathologists Cancer Protocols: Optimizing Format for Accuracy and Efficiency. Arch Pathol Lab Med. 2016;140:578–87.

5 Stogryn S, Hardy KM, Abou-Setta AM, et al. Advancement in the quality of operative documentation: A systematic review and meta-analysis of synoptic versus narrative operative reporting. The American Journal of Surgery. 2019;218:624–30.

6 E H. The Oncologist’s Guide to Synoptic Reporting: A Primer. 3298.

7 Reeves ME, Mudgway R, Lee SK, et al. Do Better Operative Reports Equal Better Surgery? A Comparative Evaluation of Compliance With Operative Standards for Cancer Surgery. Am Surg. 2020;86:1281–8.

8 Hieken TJ, Burns WR, Francescatti AB, et al. Technical Standards for Cancer Surgery: Improving Patient Care through Synoptic Operative Reporting. Ann Surg Oncol. 2022;29:6526–33.

9 Cancer Protocol Templates. College of American Pathologists. https://www.cap.org/protocols-and-guidelines/cancer-reporting-tools/cancer-protocol-templates (accessed 22 March 2024)

10 Becker G, Kempf DE, Xander CJ, et al. Four minutes for a patient, twenty seconds for a relative - an observational study at a university hospital. BMC Health Serv Res. 2010;10:94.

11 Lankshear S, Srigley J, McGowan T, et al. Standardized synoptic cancer pathology reports - so what and who cares? A population-based satisfaction survey of 970 pathologists, surgeons, and oncologists. Arch Pathol Lab Med. 2013;137:1599–602.

12 Renshaw AA, Gould EW. The Cost of Synoptic Reporting. Arch Pathol Lab Med. 2017;141:15–6.

13 Savova GK, Tseytlin E, Finan SP, et al. DeepPhe - A Natural Language Processing System for Extracting Cancer Phenotypes from Clinical Records. Cancer Res. 2017;77:e115–8.

14 Giannaris PS, Al-Taie Z, Kovalenko M, et al. Artificial Intelligence-Driven Structurization of Diagnostic Information in Free-Text Pathology Reports. Journal of Pathology Informatics. 2020;11:4.

15 Lam H, Nguyen F, Wang X, et al. An accessible, efficient, and accurate natural language processing method for extracting diagnostic data from pathology reports. J Pathol Inform. 2022;13:100154.

16 Mu Y, Tizhoosh HR, Tayebi RM, et al. A BERT model generates diagnostically relevant semantic embeddings from pathology synopses with active learning. Commun Med. 2021;1:1–13.

17 Wee-Ming Tan HSZ Kean-Hooi Teoh, Mogana Darshini Ganggayah, Nur Aishah Taib, Dhillon SK. Automated Generation of Synoptic Reports from Narrative Pathology Reports in University Malaya Medical Centre Using Natural Language Processing. 2022. 10.3390/diagnostics12040879

18 Bitterman DS, Miller TA, Mak RH, et al. Clinical Natural Language Processing for Radiation Oncology: A Review and Practical Primer. International Journal of Radiation Oncology*Biology*Physics. 2021;110:641– 55.

19 Korngiebel DM, Mooney SD. Considering the possibilities and pitfalls of Generative Pre-trained Transformer 3 (GPT-3) in healthcare delivery. npj Digit Med. 2021;4:1–3.

20 Thirunavukarasu AJ, Ting DSJ, Elangovan K, et al. Large language models in medicine. Nature Medicine. 2023;29:1930–40.

21 Goel A, Gueta A, Gilon O, et al. LLMs Accelerate Annotation for Medical Information Extraction. Proceedings of the 3rd Machine Learning for Health Symposium. PMLR 2023:82–100. https://proceedings.mlr.press/v225/goel23a.html (accessed 25 March 2024)

22 Wei J, Tay Y, Bommasani R, et al. Emergent Abilities of Large Language Models.

23 Singhal K, Azizi S, Tu T, et al. Large language models encode clinical knowledge. Nature. 2023;620:172– 80.

24 Liévin V, Hother CE, Motzfeldt AG, et al. Can large language models reason about medical questions? Patterns. 2024;5:100943.

25 Zhang T, Kishore V, Wu F, et al. BERTScore: Evaluating Text Generation with BERT. 2020. 10.48550/arXiv.1904.09675

26 Kaggal VC, Elayavilli RK, Mehrabi S, et al. Toward a Learning Health-care System – Knowledge Delivery at the Point of Care Empowered by Big Data and NLP. Biomed Inform Insights. 2016;8:13–22.

27 Geller SA, Horowitz RE. Gross examination. Methods Mol Biol. 2014;1180:3–19.

28 Wilson LB. A METHOD FOR THE RAPID PREPARATION OF FRESH TISSUES FOR THE MICROSCOPE. Journal of the American Medical Association. 1905;XLV:1737.

29 Black C, Marotti J, Zarovnaya E, et al. Critical evaluation of frozen section margins in head and neck cancer resections. Cancer. 2006;107:2792–800.

30 Sanh V, Webson A, Raffel C, et al. Multitask Prompted Training Enables Zero-Shot Task Generalization. 2022. http://arxiv.org/abs/2110.08207 (accessed 29 March 2024)

31 Devlin J, Chang M-W, Lee K, et al. BERT: Pre-training of Deep Bidirectional Transformers for Language Understanding. 2019. 10.48550/arXiv.1810.04805

32 Radford A, Wu J, Child R, et al. Language Models are Unsupervised Multitask Learners.

33 Touvron H, Martin L, Stone K, et al. Llama 2: Open Foundation and Fine-Tuned Chat Models. 2023. http://arxiv.org/abs/2307.09288 (accessed 23 March 2024)

34 Wolf T, Debut L, Sanh V, et al. HuggingFace’s Transformers: State-of-the-art Natural Language Processing. 2020. http://arxiv.org/abs/1910.03771 (accessed 29 March 2024)

35 Sushil M, Kennedy VE, Mandair D, et al. CORAL: Expert-Curated medical Oncology Reports to Advance Language Model Inference. NEJM AI. 2024;1. doi: 10.1056/AIdbp2300110

36 Zhang B, Liu Z, Cherry C, et al. When Scaling Meets LLM Finetuning: The Effect of Data, Model and Finetuning Method. 2024. http://arxiv.org/abs/2402.17193 (accessed 29 March 2024)

37 Lang H, Agrawal MN, Kim Y, et al. Co-training Improves Prompt-based Learning for Large Language Models. Proceedings of the 39th International Conference on Machine Learning. PMLR 2022:11985–2003. https://proceedings.mlr.press/v162/lang22a.html (accessed 29 March 2024)

38 Dettmers T, Pagnoni A, Holtzman A. QLORA: Efficient Finetuning of Quantized LLMs. 2305.

39 Wang R, Zelikman E, Poesia G, et al. Hypothesis Search: Inductive Reasoning with Language Models. 2023. 10.48550/arXiv.2309.05660

40 Tang X, Zheng Z, Li J, et al. Large Language Models are In-Context Semantic Reasoners rather than Symbolic Reasoners. 2023. http://arxiv.org/abs/2305.14825 (accessed 29 March 2024)

41 Zwillinger D, Kokoska S. CRC Standard Probability and Statistics Tables and Formulae. CRC Press 1999.

